# Segmentation of clinical imagery for improved epidural stimulation to address spinal cord injury

**DOI:** 10.1101/2025.06.18.25329862

**Authors:** Jordan K. Matelsky, Pawan Sharma, Erik C. Johnson, Siqi Wang, Maxwell Boakye, Claudia Angeli, Gail F. Forrest, Susan J. Harkema, Francesco Tenore

**Affiliations:** The Johns Hopkins University Applied Physics Laboratory, Laurel, MD, USA; Kessler Foundation, West Orange, NJ, USA; University of Louisville, Kentucky Spinal Cord Injury Research Center, Louisville, KY, USA; University of Pennsylvania, Department of Bioengineering, Philadelphia, PA, USA

## Abstract

Spinal cord injury (SCI) can severely impair motor and autonomic function, with long-term consequences for quality of life. Epidural stimulation has emerged as a promising intervention, offering partial recovery by activating neural circuits below the injury. To make this therapy effective in practice, precise placement of stimulation electrodes is essential — and that requires accurate segmentation of spinal cord structures in MRI data. We present a protocol for manual segmentation tailored to SCI anatomy, and evaluated a deep learning approach using a U-Net architecture to automate this segmentation process. Our approach yields accurate, efficient segmentation that identify potential electrode placement sites with high fidelity. Preliminary results suggest that this framework can accelerate SCI MRI analysis and improve planning for epidural stimulation, helping bridge the gap between advanced neurotechnologies and real-world clinical application with faster surgeries and more accurate electrode placement.

## I. Introduction

Patients with spinal cord injury (SCI) experience a wide range of symptoms, including restriction of motor, sensory, and autonomic function. This can include difficulties with cardiovascular, respiratory, and bladder function. For example, rates of orthostatic hypotension can reach 82% for people with tetraplegia and 50% for people with paraplegia immediately after injury [1]. This can result in reduced cognitive function, decreased quality of life, and increased risk of stroke [2].

Few technologies exist to address the range of functional deficits shown in SCI patients. Combinations of pharmacologic approaches and patient education can improve orthostatic hypotension symptoms [3]. However, a common recommendation for people with orthostatic hypotension and SCI is to simply adapt to fluctuations in blood pressure, as the damage to spinal cord sympathetic neurons limits potential for improvements [3], [4]. This is further complicated by the fact that multiple targets (e.g., blood pressure, bladder function, and motor control) must be regulated simultaneously.

To address the limitations in current interventions, there is ongoing research into the utilization of spinal cord epidural stimulation (scES), a process in which the spinal cord is electrically stimulated via a surgically implanted electrode grid [5]. Epidural stimulation can regulate systolic blood pressure in people with SCI and chronic orthostatic hypotension [4], [6]. In addition, stimulation can result in adaptation of neural circuitry to improve outcomes and has the potential to influence other functions such as bladder control and voluntary motor control [7], [8].

Critical to determining outcomes and effectiveness of epidural stimulation is individualized placement of electrodes. This requires accurate presurgical analysis of anatomical imaging results, for example from Magnetic Resonance Imaging (MRI). Moreover, this analysis needs to be carefully integrated into workflows to help neurosurgeons guide electrode placement during electrode placement surgery. Recent work has focused on collecting data to assess these kinds of placement problems [9], [10], representing an opportunity for analysis and visualization tools and pipelines.

Directly interpreting 2D MRI slices during surgical planning is challenging — especially when trying to reason about the 3D spatial relationships between electrodes and critical spinal anatomy. Planning electrode placement requires a mental reconstruction of complex geometry, which is error-prone and inefficient. To address this, we need a pipeline that converts raw MRI data into interactive 3D visualizations of key neuroanatomical landmarks. This involves collecting highquality imaging, manually segmenting structures of interest (a time-intensive process requiring anatomical expertise), and implementing strategies to reduce inter-observer variability. When combined with lightweight 3D rendering tools, this workflow could offer intuitive, spatially accurate visualizations that support surgical decision-making and improve outcomes in SCI patients.

To address the challenges and rate-limiters of this workflow — particularly the manual annotation of key anatomical structures — preliminary automated image segmentation methods have been developed. A variety of architectures have arisen in this space: One such method is the U-Net architecture, which has been successfully applied to a variety of medical imaging tasks [11]. A second and more recent architecture that has grown in popularity is the Vision Transformer, an architecture that uses *attention* in pixel-space to perform context-aware learning on an image [12], [13].

In this study, we develop a pipeline for 3D visualization to aid spinal cord epidural stimulation electrode placement. We also present preliminary use of deep learning techniques to automated image segmentation of MRI data for SCI clinical data. We share evaluation results for surgical electrode placement, and find improved electrode placements while decreasing the amount of time patients spend in surgery. Further automation methods may allow widespread deployment, though more work is required to validate and generalize the computer vision segmentation in the SCI population.

## II. Methods

Large and high-dimension image visualization is a common challenge for bioimagery analysis. We have developed a novel visualization pipeline that converts expert-segmented Magnetic Resonance Imaging (MRI) data into 3D data which is explorable in real-time. This provides low-latency and interactive visualizations to clinicians for exploration in a common web browser on consumer hardware (**Fig. 1**). These visualizations enable more intuitive and navigable environments for clinician review of patient anatomy prior to surgical electrode placement. Annotations can be created through manual image segmentation or a trained machine learning segmentation algorithm. We utilize this pipeline to leverage existing segmentation data from the visualization stage in order to bootstrap a domain specific training dataset for SCI segmentation.

**Fig. 1.**
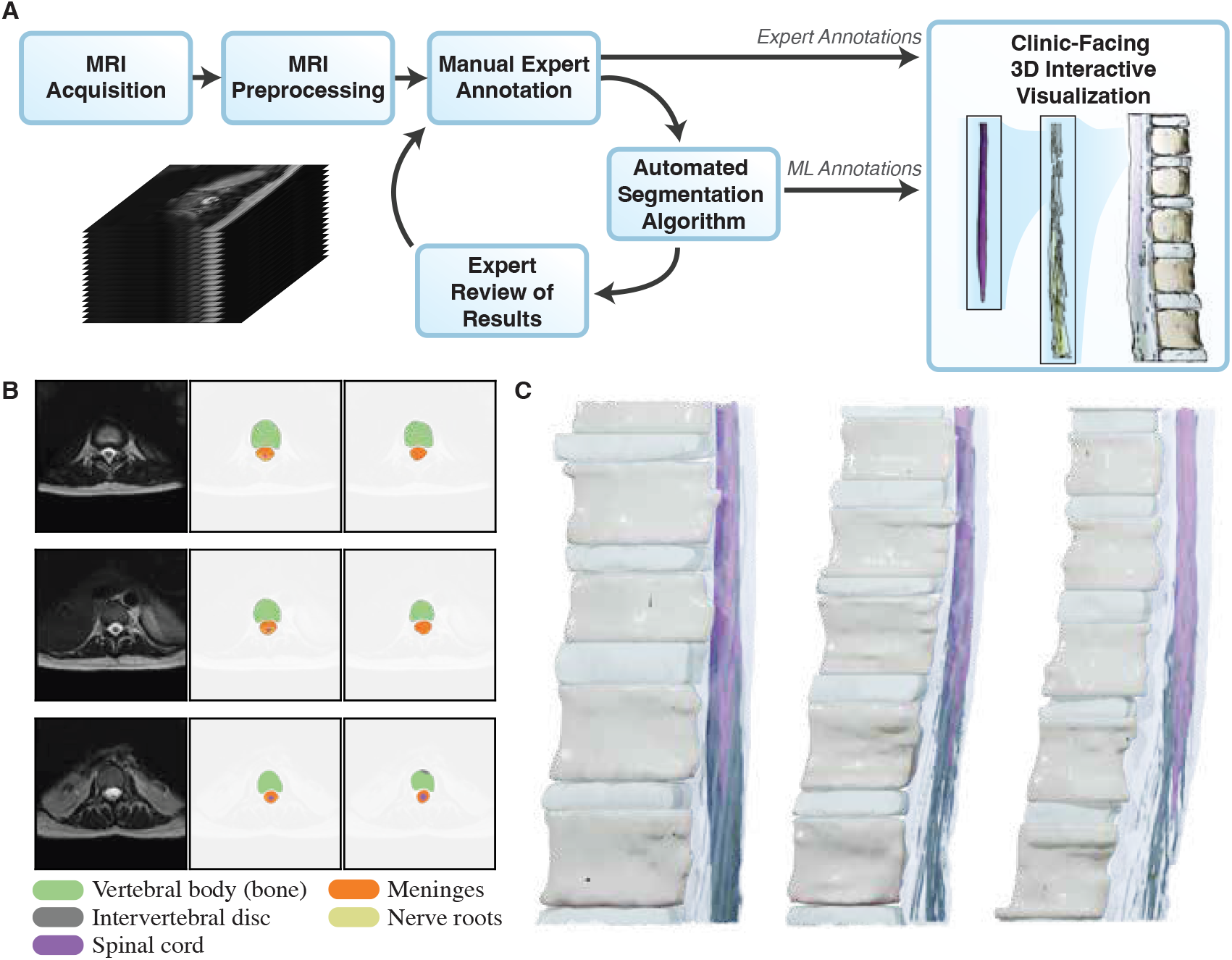
Overview of the 3D visualization approach, automated segmentation, and example 3D renderings. **A)** The segmentation pipeline, progressing from an MRI image stack through preprocessing. At this stage, manual annotations of 2D slices can be created by expert neuroanatomical annotators. This results in expert annotations which can be visualized in 3D. In addition, automated algorithms can be trained using the resulting manual annotations (with expert review). **B)** Examples from three slices of an MRI image (first column), the ground truth annotation labels (second column) and output of the automated U-net approach (third column). **C)** Example 3D reconstructions for three participants, which can be provided as an interactive visualization for clinicians.

### A. Participants

50 individuals (32 Males, 18 Females, 9.0±8.1years since injury) diagnosed with chronic motor spinal cord injury (SCI) were enrolled to participate in the studies investigating motor rehabilitative effects of activity-based training combined with spinal cord epidural stimulation (scES). All research participants were over 19 years (34.0±9.5 years) of age at the time of scES implant and met the following inclusion criteria: non-progressive SCI at the cervical and thoracic spinal cord (C3-T5, AIS A, B or C) and at least six months postinjury and with no medical conditions unrelated to SCI. Participants were either implanted with the Medtronic RestoreAdvanced™ stimulation (voltage-based stimulation) or Medtronic Intellis™ stimulation (current-based stimulation) system. Both Medtronic systems use 16-electrode Specify^®^ 5-6-5 leads. scES implantation surgery time was recorded for all participants. All participants provided written informed consent before participating in the study. The experimental protocol was approved by the University of Louisville Institutional Review Board and was in accordance with the Declaration of Helsinki. 25 participants underwent spinal cord neuroanatomical mapping prior to implant surgery, and the remaining 25 participants post-implant surgery. From each group, lumbosacral enlargement coverage by scES paddle analysis was performed for 19 participants.

### B. Spinal cord neuroanatomical mapping

Neuroanatomical mapping was performed using an established and well-standardized methodology [14], [15]. Briefly, high-resolution axial and sagittal MRI scans (3 mm slice thickness and zero gaps) of the lumbar and sacral spinal cord were obtained to locate and manually annotate the dorsal roots, ventral roots, signal cord segments, and vertebrae. The location and annotation of the roots, segments, and vertebrae were identified using RadiAnt DICOM Viewer™ and marked using both Mango (Multi-image Analysis GUI) and 3D Slicer (software to visualize and process medical images). As the spinal cord typically ends at the L1 vertebral level, the lumbar and sacral nerve roots exit the spinal canal further distally compared to the corresponding spinal cord segment giving rise to these roots (L1, L2, …, S1). The lumbar (L1-L5) and sacral (S) spinal cord segments were anatomically estimated by identifying the dorsal and ventral roots on the left and right sides that exit the spinal canal at each vertebral level and back-tracing those nerve roots into the spinal cord body. A 3D model of the spinal cord of each individual was then reconstructed based on the obtained annotations using custom-written software (see **II-C**). To identify the location of the ES paddle on the spinal cord, anterior-posterior and lateral X-ray images of the spinal cord were obtained. The anterior-posterior image was used to identify the T12 vertebra, and the lateral image was used to locate the rostral and caudal ends of the paddle in relation to the vertebral bodies. Later, the location of the paddle over the spinal cord was estimated by integrating the lateral X-ray with the sagittal and axial MRI scans. Based on the length of the paddle electrode (46.5 mm for Medtronic Specify^®^ 5-6-5 lead), 15 consecutive MRI axial slices (total of 15 × 3 mm = 45 mm in length) were identified that best describe its location over the cord. The paddle electrode was superimposed over the identified 15 images in the 3D model to obtain the percentage of volumetric coverage of the lumbosacral spinal cord by the scES paddle electrode for each participant. All annotations were performed by an expert and trained analyst and further checked by a neuroradiologist on the team for accuracy. Additionally, Cohen’s Kappa was calculated to assess inter-rater reliability. The analysis showed a kappa value between 0.8 and 0.9, indicating substantial agreement and consistency between raters while annotating different structures.

### C. 3D Visualization

Slice-based viewers and annotation engines are unsurpassed for generating dense segmentation of 3D volumes, but tend to provide less user-friendly spatial intuitions about the data. In order to convert segmentation data into interpretable renders, we converted the segmentation channels from 2D slices into a single 3D volume in Python using the *Pillow* [16] and *numpy* [17] libraries. We then converted this set of binary masks to a sparse mesh representation using a bespoke marching cubes algorithm derived from the *PyMCubes* library [18]. Performant high-dimension image visualization is a common challenge for bioimagery analysis. While software written to run natively on the operating system tends to have great performance advantages, web-based visualization engines are powerful tools for accessibility and reproducibility [19]. We leveraged *Neuroglancer* [20], an existing 3D visualization tool, in order to deliver low-latency and interactive visualizations of these meshes and image channels to clinicians for exploration. These segmentations can also be exported for rendering in traditional 3D rendering software. The visualizations included in **Fig. 1** and **2** were rendered with the EEVEE realtime rendering engine in Blender. [21]

### D. Preliminary Automated Segmentation

Semantic segmentation is an increasingly routine component of the bioimagery machine learning toolkit. Previous work established that U-Nets — a symmetric downsample/upsample architecture — are powerful and highly performant models for segmenting dense 3D biological tissue volumes [22], [23] with much lower training-data volume requirements than the more recent Vision Transformer architectures [12], [13]. Training data volume is a key challenge here due to the manual labor requirements of hand-segmentation, and because SCI has different visual and anatomical properties that make cross-training across spinal MRI datasets nontrivial. Adopting these lessons, we developed a pipeline to leverage existing segmentation efforts during the visualization stage in order to bootstrap a domain-specific training dataset for SCI segmentation.

We trained a U-net model to minimize loss on a dataset consisting of 42 3D volumes hand-verified for quality control. Eight volumes were reserved for hold-out assessment. Data volumes were augmented through axial/yaw 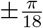 rotation. along the vertical axis (i.e., parallel to the spinal cord) 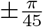 and rotations along the mediolateral axis (i.e., forward/backward). Rotation angles were sampled uniformly. Further augmentation applied pixelwise Gaussian noise of 𝒩(0, 0.05) to the normalized pixel intensities.

A 3D U-net architecture was created with four down-sampling and four upsampling blocks, with a single image channel to represent MRI greyscale data in uint16 bitdepth, and 14 initial convolution filters. Volumes were resized to 48 *×* 512 *×* 512 (ZYX) voxels for training and testing. Models were trained using PyTorch Lightning for a maximum of 100 epochs using Adam optimization with a grid-search learning rate of *η* = 1.2 *×* 10^*−*3^. After training, we evaluated the performance of the model on the held-out test set of spinal cord MRI images. We report the results in terms of mean intersection-over-union (mIoU) and pixel accuracy.

### E. Assessment for Surgical Placement

This technique improves the ability to plan surgical placement of the stimulating electrode paddle. To evaluate the impact of this new visualization technique, the neurosurgical team is provided with pre-surgical imaging (MRIs, CTs) and are asked to indicate on the image the location where they would place the electrodes. After this, we provide them with the 3D visualization and ask them to do the same task, and we compute the distance between the two locations. After the surgery, we solicit and incorporate neurosurgeon feedback into updates to the visualization and segmentation tools.

Models of the 3D reconstruction of the lumbosacral spinal cord, showing the relationship between the spinal cord and the vertebrae for each participant, are provided to the surgeon. Several potential placements are proposed by our team, with the electrode lead projected onto the vertebrae corresponding to a participant’s lumbosacral spinal cord. These representations are confirmed with the fluoroscopy images taken during the surgical procedure.

## III. Results

Our visualization pipeline has been developed and applied to patient MRI data to create visualizations for use in surgical planning for 50 participants. This automated segmentation approach can be used for data segmentation or trusted segmentations can be generated by manual annotators. Datasets for training automated algortihms are being improved over time and can be refined with manual input as needed. Three dimensional visualizations are then provided for planning of electrode placement. This workflow is captured in **Fig. 1A**, where data move from acquisition to image stacks to segmentation to 3D rendering, ultimately producing visualizations for surgical planning.

Examples of the 3D visualization pipeline can be seen in **Fig. 1C**, which captures key steps in the workflow. **Fig. 1B**, left column, shows example 2D MRI images, without labeling of key anatomical features for the spinal cord. This represents the raw data available for surgical planing. **Figs. 1B**, center and right columns, show the expert manual annotation and automated annotations for these slices. Key anatomical regions such as the vertebral column, spinal cord, and nerve fibers are labeled and visualized with different colors. After segmentation of a 2D volume stack, a 3D mesh can be generated (**Fig. 1C**). Finally, a 3D visualization, including anatomical labels, can be created for final surgical planning and provided as a 3D, interactive visualization using our system.

**Figure 2A** presents the results of this process from patient MRI data using manual expert segmentations. We have completed *N* = 25 individuals using the lumbosacral 3D reconstruction to inform the neurosurgeon for placement decisions. For *N* = 19 of those individuals, post-surgical lumbosacral enlargement volume and length covered by scES paddle were estimated to assess placement quality. Feedback from the neurosurgeon indicated that our approach provides “good visualization of the lumbosacral enlargement of the conus — superimposed paddle on the MRI helps develop amount of coverage or percentage of the lumbosacral enlargement covers by the stimulator [grid] — this in turn helps guide the entry side for the laminotomy.” The precision of the placement influences the outcomes as quantified by the relationship between ability of clinically motor complete SCI individuals to move their legs voluntarily and the accuracy of the placement of the electrode lead over the lumbosacral spinal cord as quantified by percentage of lumbosacral enlargement volume covered by scES (see **Fig. 2A**). Those individuals who had the 3D reconstruction available for surgery had a significantly higher percentage of coverage (*p* < 0.001) of the lumbosacral spinal cord than those who did not (**Fig. 2C**). We also compared the surgical times from these individuals to those we had implanted before this was available and found significantly shorter surgery times when using our 3D reconstruction (*p* = 0.013, see **Figure 2B**).

**Fig. 2.**
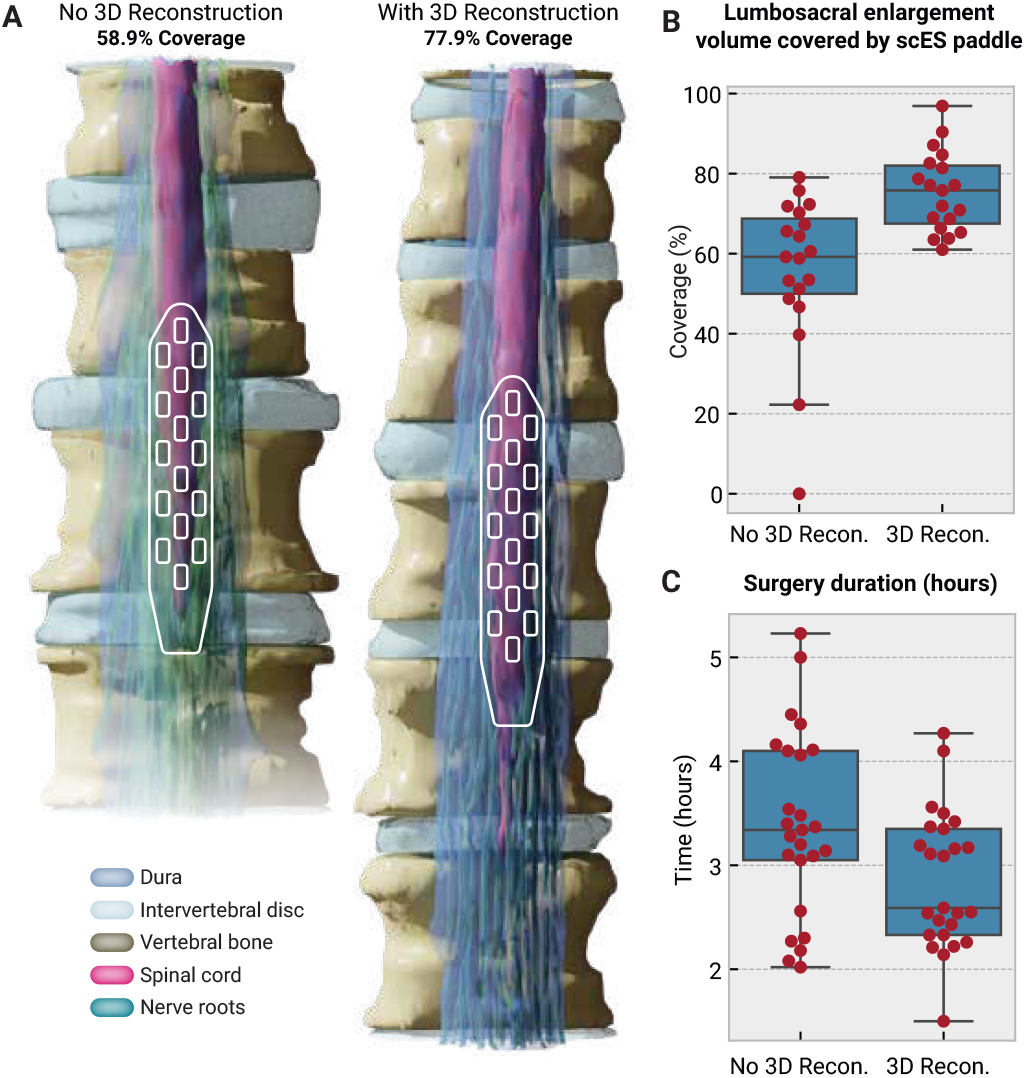
Lumbosacral cord 3D reconstruction, associated measurements and surgery time. **A)** Shown are the 3D reconstructed images of lumbosacral enlargement with scES paddle electrode for two representative participants that underwent 3D reconstruction of the spinal cord after (manual) and prior to (automated) scES surgery. **B)** Box plots show lumbosacral enlargement volume length covered by scES paddle electrode. (Higher is better.) **C)** Box plots for surgery time in manual and automated reconstruction groups. (Lower is better.)

As manual annotation can be time consuming and require expert neuroanatomical knowledge, we have utilized our training dataset to train a U-net segmentation approach to help accelerate future surgical planning. **Figure 3** shows the crossentropy loss for training the automated segmentation approach, demonstrating a clear decrease in both training and validation loss. **Figure 1B** shows the output of this approach. The automated segmentation approach, while efficient, does still have high error for certain anatomical structures, such as nerve roots, which are individually smaller than the voxel resolution of the MRI scan and have been historically found to be difficult to segment through convolutional neural networks. Final mean Intersection over Union was calculated as 0.58 and 0.44 for the training and validation sets, respectively. This suggests there is room for improvement, particularly on high-error classes such as nerve fibers. However, more macroscale structures are well identified.

**Fig. 3.**
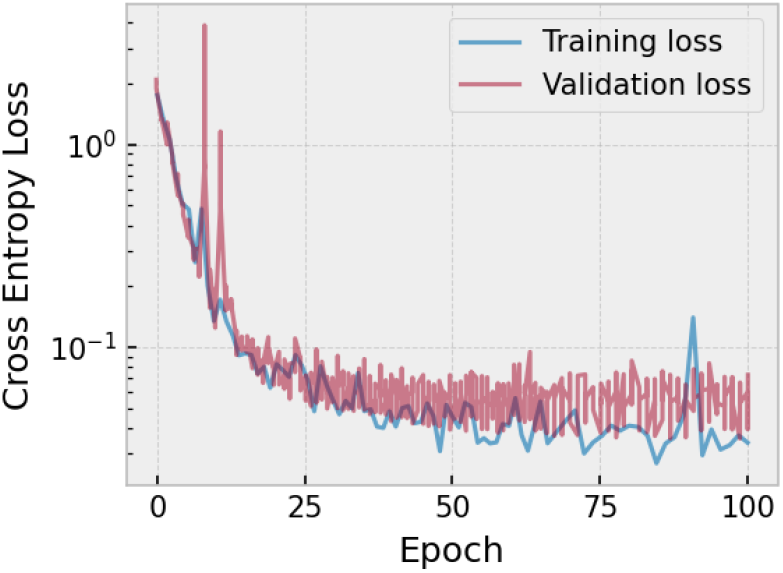
Cross-entropy loss curve for training the U-net approach. Training and validation curves are both shown. There is a small gap between training and validation loss, suggesting overfitting is not a dominant issue.

## IV. Discussion and Conclusions

We have presented a pipeline for 3D visualization and analysis to guide surgical placement of scES electrodes, which can be used to improve surgical outcomes. In particular, our results demonstrate reduction in surgery time and improvements in electrode coverage that can translate directly into improved outcomes. This work improves over our previous efforts [5] through creation of this semi-automated pipeline and testing in surgical conditions. There may be implications of this approach for other surgical procedures requiring precise placement of electrodes relative to spinal cord tissue.

One aspect of this pipeline development process is creating a reproducible workflow which can be utilized at a range of surgical centers. Many aspects, such as mesh generation, 3D rendering, and the interactive web visualization are based on open-source tools which can be rapidly adapted to different scanners and use cases. This permits easy deployment in new surgical settings. Annotation, whether manual or automated, however, requires training of human experts and/or fine tuning of segmentation approaches. Further work will be required to streamline the annotation approach for new surgical centers.

The automated segmentation approach remains an area for further work, as outputs have not yet reached the accuracy of manual segmentation. In particular, segmentation of the fine nerve fiber structures in 3D is challenging from 2D or shallow 3D U-nets. Two approaches may be possible to overcome this in the future. First, continuing to develop datasets for segmentation (e.g. [9], [10]) with sufficient individual and scanner variability may help create a more generalizable approach. Second, 3D segmentation approaches which weight spatial continuity of the nerve fibers through the volume, incorporating biological priors on nerve fiber length and distribution, may reduce 3D segmentation errors of these fine structures. Moreover, work is required to further develop a fine-tuning pipeline to adapt pre-trained weights to particular scanners and surgical settings, through collecting small amounts of expertly annotated data to update the model. In addition, procedures will be required to assess generalization performance to ensure automated reconstructions are of sufficient quality to assist surgical planning. Future work will investigate the use of the automated segmentations for improving surgical outcomes.

Overall, we have demonstrated a semi-automated pipeline for 3D segmentation and visualization of MRI data to assist surgical placement of scES electrodes. Given the implications and improvements in surgical time and electrode placement, this method should be further explored for new surgical sites. Further work will be required to improve packaging for deployment and automation of segmentation strategies.

## Data Availability

The data are not publicly available due to privacy or ethical restrictions.

## V. Acknowledgements

We thank the participants for their courage, dedication, motivation, and perseverance that made developing this system possible; the University of Louisville and Kessler Foundation research staff for support. We would also like to thank Brock Wester and Teresa Colella for helping foster and guide this work.

This work was made possible, in part, through financial support from the Kessler Foundation, a University of Louisville grant (2016PG-MED001, Leona M. & Harry B. Helmsley Charitable Trust; ES BI-2017; University of Louisville Hospital; Medtronic Plc) and internal JHU/APL research funding. This work was also supported by the NIH Neuromodulation Prize. The views, opinions and/or findings expressed are those of the authors and should not be interpreted as representing the official views or policies of the U.S. Government.

## References

[1] A Illman, K Stiller, and M Williams. The prevalence of orthostatic hypotension during physiotherapy treatment in patients with an acute spinal cord injury. Spinal Cord, 38(12):741–747, 2000.

[2] Patricia Branco Mills, Carlen K Fung, Angelique Travlos, and Andrei Krassioukov. Nonpharmacologic management of orthostatic hypotension: a systematic review. Archives of physical medicine and rehabilitation, 96(2):366–375, 2015.

[3] Andrei Krassioukov, Janice J Eng, Darren E Warburton, Robert Teasell, Spinal Cord Injury Rehabilitation Evidence Research Team, et al. A systematic review of the management of orthostatic hypotension after spinal cord injury. Archives of physical medicine and rehabilitation, 90(5):876–885, 2009.

[4] Bonnie E Legg Ditterline, Sevda C Aslan, Siqi Wang, Beatrice Ugiliweneza, Glenn A Hirsch, Jill M Wecht, and Susan J Harkema. Restoration of autonomic cardiovascular regulation in spinal cord injury with epidural stimulation: a case series. Clinical Autonomic Research, 31(2):317–320, 2021.

[5] Erik C Johnson, Jordan K Matelsky, Christa Cooke, Breanne Christie, Khalid Jones, Harley Ledbetter, Siqi Wang, Gail Forrest, Nathan Torgerson, Claudia A Angeli, et al. Automated tools to improve spinal cord injury outcomes with epidural stimulation. In 2023 11th International IEEE/EMBS Conference on Neural Engineering (NER), pages 1–4. IEEE, 2023.

[6] Susan J Harkema, Siqi Wang, Claudia A Angeli, Yangsheng Chen, Maxwell Boakye, Beatrice Ugiliweneza, and Glenn A Hirsch. Normalization of blood pressure with spinal cord epidural stimulation after severe spinal cord injury. Frontiers in human neuroscience, page 83, 2018.

[7] Susan J Harkema, Yury Gerasimenko, Jonathan Hodes, Joel Burdick, Claudia A Angeli, Yangsheng Chen, Christie Ferreira, Andrea Willhite, Enrico Rejc, Robert G Grossman, et al. Effect of epidural stimulation of the lumbosacral spinal cord on voluntary movement, standing, and assisted stepping after motor complete paraplegia: a case study. The Lancet, 377(9781):1938–1947, 2011.

[8] Nathan Greiner, Beatrice Barra, Giuseppe Schiavone, Henri Lorach, Nicholas James, Sara Conti, Melanie Kaeser, Florian Fallegger, Simon Borgognon, Stéphanie Lacour, et al. Recruitment of upper-limb motoneurons with epidural electrical stimulation of the cervical spinal cord. Nature communications, 12(1):1–19, 2021.

[9] Jionghui Liu, Wenqi Zhang, Yuxing Zhou, Linhao Xu, Ying-Hua Chu, and Fumin Jia. An open-access lumbosacral spine mri dataset with enhanced spinal nerve root structure resolution. Scientific Data, 11(1):1131, 2024.

[10] Julien Cohen-Adad, Eva Alonso-Ortiz, Mihael Abramovic, Carina Arneitz, Nicole Atcheson, Laura Barlow, Robert L Barry, Markus Barth, Marco Battiston, Christian Büchel, et al. Open-access quantitative mri data of the spinal cord and reproducibility across participants, sites and manufacturers. Scientific Data, 8(1):219, 2021.

[11] Olaf Ronneberger, Philipp Fischer, and Thomas Brox. U-net: Convolutional networks for biomedical image segmentation, 2015. cite arxiv:1505.04597Comment: conditionally accepted at MICCAI 2015.

[12] Bingzhi Chen, Yishu Liu, Zheng Zhang, Guangming Lu, and Adams Wai Kin Kong. Transattunet: Multi-level attention-guided u-net with transformer for medical image segmentation. arXiv preprint arXiv:2107.05274, 2021.

[13] Abhinav Sagar. Vitbis: Vision transformer for biomedical image segmentation, 2022.

[14] Samineh Mesbah, Tyler Ball, Claudia Angeli, Enrico Rejc, Nicholas Dietz, Beatrice Ugiliweneza, Maxwell Boakye, and Susan Harkema. Predictors of volitional motor recovery with epidural stimulation in individuals with chronic spinal cord injury. Brain, 144(2):420–433, 2021.

[15] Samineh Mesbah, April Herrity, Beatrice Ugiliweneza, Claudia Angeli, Yuri Gerasimenko, Maxwell Boakye, and Susan Harkema. Neuroanatomical mapping of the lumbosacral spinal cord in individuals with chronic spinal cord injury. Brain Communications, 5(1):fcac330, 2023.

[16] Alex Clark. Pillow (pil fork) documentation, 2015.

[17] Charles R. Harris, K. Jarrod Millman, Stéfan J van der Walt, Ralf Gommers, Pauli Virtanen, David Cournapeau, Eric Wieser, Julian Taylor, Sebastian Berg, Nathaniel J. Smith, Robert Kern, Matti Picus, Stephan Hoyer, Marten H. van Kerkwijk, Matthew Brett, Allan Haldane, Jaime Fernández del Río, Mark Wiebe, Pearu Peterson, Pierre Gérard-Marchant, Kevin Sheppard, Tyler Reddy, Warren Weckesser, Hameer Abbasi, Christoph Gohlke, and Travis E. Oliphant. Array programming with NumPy. Nature, 585:357–362, 2020.

[18] @pmneila. Pymcubes. github.com/pmneila/PyMCubes, 2020.

[19] Jordan K Matelsky, Joseph Downs, Hannah P Cowley, Brock Wester, and William Gray-Roncal. A substrate for modular, extensible data-visualization. Big data analytics, 5(1):1–15, 2020.

[20] Jeremy Maitin-Shepard et al. Neuroglancer. github.com/google/neuroglancer, 2021.

[21] Blender Online Community. Blender - a 3d modelling and rendering package, 2018.

[22] Özgün Ç iÇek, Ahmed Abdulkadir, Soeren S Lienkamp, Thomas Brox, and Olaf Ronneberger. 3D U-Net: learning dense volumetric segmentation from sparse annotation. In International conference on medical image computing and computer-assisted intervention, pages 424–432. Springer, 2016.

[23] Yang Li, Qianqian Yao, Haitao Yu, Xiaofeng Xie, Zeren Shi, Shanshan Li, Hui Qiu, Changqin Li, and Jian Qin. Automated segmentation of vertebral cortex with 3D U-Net-based deep convolutional neural network. Frontiers in Bioengineering and Biotechnology, page 1753.

